# Effects of front-of-package nutrition labels in Latine and limited English proficiency populations: A randomized trial

**DOI:** 10.1101/2025.05.09.25327177

**Authors:** Marissa G. Hall, Cristina J. Y. Lee, Aline D’Angelo Campos, Natalicio Serrano, Lindsey Smith Taillie, Jennifer Falbe, Aviva Musicus, Callie Whitesell, Angela Viviana Martinez, Anna H. Grummon

## Abstract

**Introduction:** The effects of front-of-package nutrition labels among Latino and Hispanic (“Latine”) adults in the US, including those with limited English proficiency, remains largely unknown. We examined the impact of different types of labels among Latine consumers and whether effects differed by English proficiency.

**Study design:** Online randomized trial.

**Setting/participants:** 3,053 Latine US adults (49% limited English proficiency).

**Intervention:** Participants viewed one of three labels: *numerical labels* displaying numerical information about sodium, saturated fat, and added sugar; *text high-in labels* stating when foods are high in these nutrients of concern; and *icon high-in labels* identical to the text labels plus a magnifying glass icon.

**Main outcome measures:** Participants viewed three frozen pies, three frozen pizzas, and three frozen meals displaying randomly assigned labels and identified the healthiest and least healthy product within each group (based on nutrient content).

**Results:** Text high-in labels (49% correct) led to higher correct identification of the least healthy foods compared to the numerical labels (44%, *p*<.001) though the icon high-in labels did not (47%, *p*=.07). Neither the text high-in labels (46% correct) nor the icon high-in labels (46%) led to better identification of the healthiest food compared to the numerical labels (45%, all *p*>=.71). Neither type of high-in label led to more correct identification of foods high in nutrients of concern or higher selection of the healthiest food for purchase compared to the numerical labels (all *p*>=.09). English proficiency moderated the impact of label type on correct identification of the least healthy food (*p*-interaction=.003) such that the benefit of high-in labels was only present for participants with high English proficiency.

**Conclusions:** High-in labels helped Latine consumers identify unhealthy foods more than numerical labels, but only among those with high English proficiency. Future food labeling research should include participants with limited English proficiency.

**Trial registration:** NCT06293963.

## Introduction

Adults in the US consume substantially more added sugars, saturated fat, and sodium than recommended,^1,2^ contributing to diet-related diseases such as obesity, type 2 diabetes, and heart disease.^3^ Latino and Hispanic (hereinafter “Latine”) populations and people with limited English proficiency are more likely to have several diet-related diseases than non-Latine-White and higher-English-proficiency populations.^4–9^ Interventions and policies to reduce diet-related diseases in these populations are urgently needed.

Front-of-package nutrition labels are a promising policy option for reducing the burden of diet-related disease. In the US, the only nutrition label required on food packaging is the Nutrition Facts Label, located on the back or side of product packaging. However, most US consumers do not use the Nutrition Facts Label frequently.^10,11^ In January 2025, the US Food and Drug Administration (FDA) released a proposed rule that would require nutrition labels on the front of all packaged foods in the US.^12^ If the rule is finalized, the US would join the nine countries in the Americas that require front-of-package nutrition labels.^13^ Research indicates that well-designed front-of-package nutrition labels can improve consumer understanding of product healthfulness and shift consumers toward healthier purchases.^14–20^ What remains unknown is how to design new front-of-package nutrition labels to ensure these benefits reach Latine and limited English proficiency populations, who comprise 19%^21^ and 8%^22^ of the US population, respectively.

Interpretative “high-in” labels that signal when foods are high in nutrients of concern (i.e., sodium, saturated fat, or added sugars exceed 20% of the daily value^23^) are an especially promising type of food label. Quasi-experimental studies suggest that high-in labels implemented in Latin American countries are associated with lower purchasing and consumption of nutrients of concern.^19,20,24^ In experimental studies, Latin-American-style high-in labels have led to better consumer understanding of product healthfulness and healthier purchases.^25–29^ High-in labels stand in contrast to non-interpretive “numerical” labels that present amounts of these nutrients in the product. Numerical labels are used voluntarily in the US.^30^ However, research suggests that many consumers struggle to understand the types of quantitative information present on these labels.^31–34^ Additional research comparing high-in to numerical labels could inform future labeling policy in the US and globally.

Another important research gap is understanding which kinds of front-of-package nutrition labels work best in Latine and limited English proficiency populations in the US, given higher rates of diet-related diseases in these groups.^4–9^ Most prior studies of nutrition labels in the US have included only general samples of English-speaking adults. Few studies have tested high-in labels among those with limited English proficiency, who may especially benefit from certain label designs (e.g., those with icons^35^), but limited research has examined this. This study aimed to determine which type of front-of-package nutrition label is most effective at helping Latine consumers identify less healthy and healthier products. The study also aimed to explore whether the effects of front-of-package nutrition labels differ by English proficiency.

## Methods

### Participants

In August and September 2024, we recruited 3,053 US adults ages 18-55 years who self-identified as Latino/a or Hispanic. To recruit the sample, we partnered with ThinkNow, a panel company specializing in market research with Latine populations in the US. Their panel includes ∼130,000 active members, with an average of 3,000+ members added each month. ThinkNow used quotas to ensure that approximately 50% of the sample had limited English proficiency, defined as reporting speaking English “not at all” or “not well” (vs. “well” or “very well”) using a measure adapted from the US Census.^22^ The University of North Carolina Institutional Review Board approved this study (#24-0300).

### Stimuli

We developed three types of front-of-package nutrition labels: a numerical label, a text high-in label, and a text high-in label with an icon (i.e., icon high-in label; **Figure 1**). The numerical label was a Guideline Daily Amounts label that listed nutrient quantities and percent daily values for added sugars, saturated fat, and sodium. FDA tested this label in 2023 when vetting potential labels.^36^ The two high-in labels signaled when products were high in added sugars, saturated fat, or sodium (defined as having >20% daily value per serving).^23^ The text high-in label was identical to one that FDA tested when vetting potential labels.^36^ The icon high-in label was identical to the text high-in label with the addition of a magnifying glass icon. We chose a magnifying glass icon because Canada’s and Brazil’s front-of-package nutrition labels include this icon.^37^

**Figure 1.**
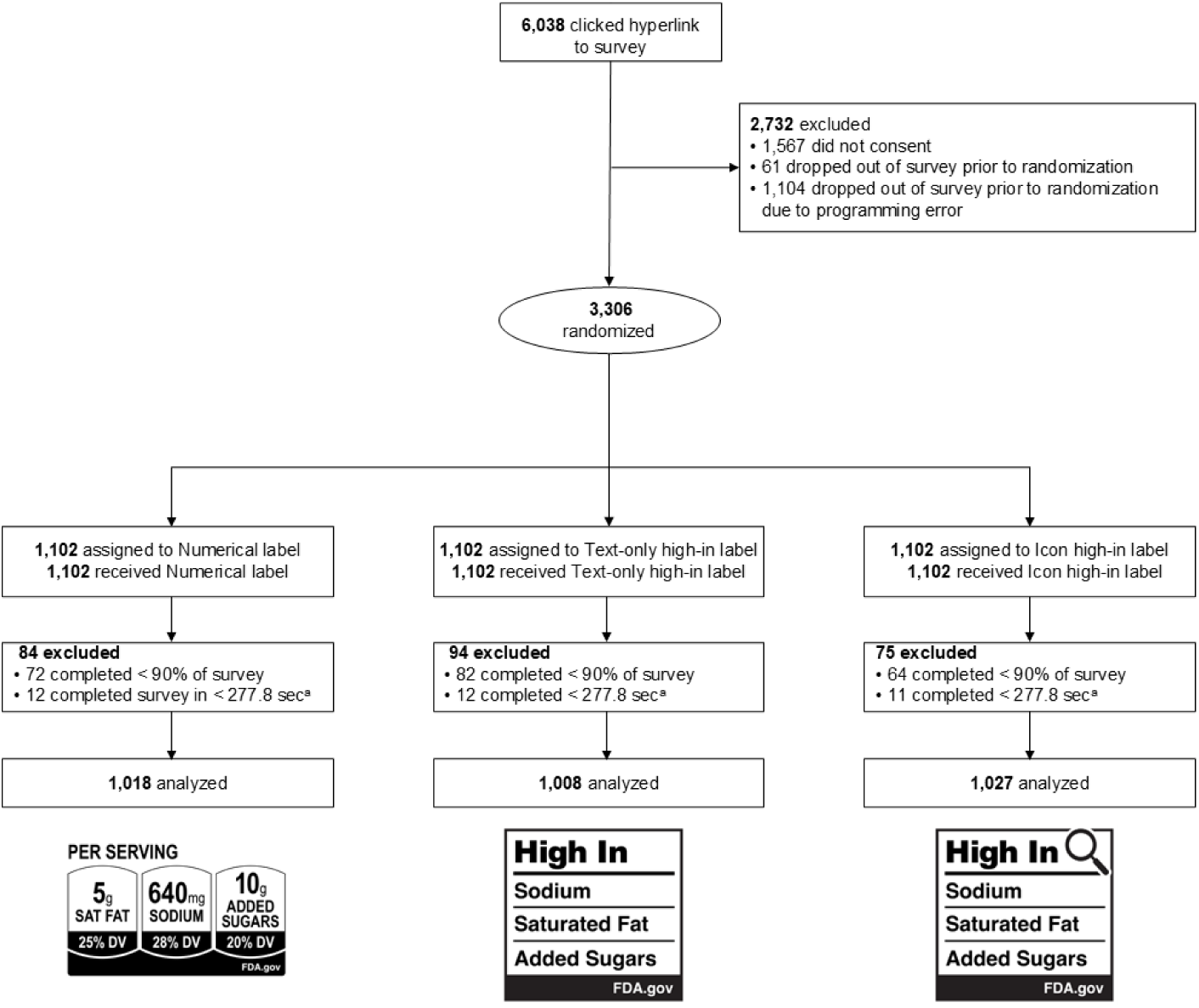
CONSORT diagram

We created images of nine different frozen foods, including three products within each of three categories: barbecue chicken pizza, sweet and sour chicken dinner, and caramel pecan pie. We selected these categories because they are plausibly high in up to three nutrients of concern (i.e., added sugars, saturated fat, and sodium). We selected the quantities of added sugar, saturated fat, and sodium in the mock products based on real foods in consultation with a registered dietician. A professional designer created study stimuli. We used fictitious branding to avoid the strong influence of established brand preferences.^38^ To heighten realism, the images displayed standard design and product information features including a picture of the food, content weight, and a description of the food. In each set, there was one product high in one nutrient of concern (i.e., the “healthiest” option), one product high in two nutrients of concern, and one product high in three nutrients of concern (i.e., the “least healthy” option).

### Procedures

After providing electronic informed consent, participants took an online survey (average length of 14 minutes), programmed in Qualtrics survey software. The trial used a between-subjects design in which participants were randomized to one of three front-of-package label conditions in a 1:1:1 allocation ratio using the Qualtrics randomizer function, with the “evenly present elements” option selected. In the survey, participants viewed all three categories of foods (displayed in random order) and completed four identification or selection tasks within each food category, described further below in *Measures*. All foods displayed participants’ randomly assigned label type. The survey did not provide any instructions or background information about the labels and did not call attention to the labels through enlargement or other methods. See **Supplementary Figure 1** for all stimuli used in the study. The font size in the numerical labels was slightly smaller than the high-in labels to mirror how the labels would be likely to be implemented in the real world. Participants then completed a series of unrelated experimental tasks and answered survey questions about demographics and other participant characteristics. After completing the survey, participants received incentives (e.g., Amazon vouchers, PayPal payments) in an amount agreed upon with ThinkNow. The survey and consent form were available in both English and Spanish; participants could select which language they preferred. These materials were translated by a professional translation company and reviewed for accuracy by four native Spanish speakers. Study stimuli were only in English.

### Measures

Exact item wording for study measures appears in **Supplementary Table 1**. The survey assessed four outcomes: correct identification of the least healthy food (co-primary outcome), correct identification of the healthiest food (co-primary outcome), correct identification of foods high in nutrients of concern (secondary outcome), and selection of the healthiest food for purchase (secondary outcome). These outcomes were measured three times for each participant: once for each of the three groups of three products. We selected the co-primary outcomes to mirror FDA’s stated goal of front-of-package nutrition labels, to help consumers “quickly and easily identify foods that can help them build a healthy eating pattern.”^12^ In terms of survey order, we measured selection first, followed by correct identification of healthiest food, least healthy food, and high in nutrients.

We measured correct identification of the least healthy food with the item: “Which of these products is the <u>least healthy</u>?” The choices were the three foods displayed in random order; participants could only select one food. Correct identification of the least healthy food was defined as selecting the food that was high in all three nutrients of concern as the least healthy.

We assessed correct identification of the healthiest food with the following item: “Which of these products is the <u>healthiest</u>?” As with the prior item, the choices were the three foods displayed in random order; participants could only select one food. Correct identification of the healthiest product was defined as selecting the food that was high in only one nutrient of concern as the healthiest.

We assessed correct identification of foods high in nutrients with the item: “Which of these products are <u>high in [nutrient]</u>?” To reduce survey burden, participants were randomly assigned to either view added sugar, saturated fat, or sodium in the question stem. Correct identification of products high in the nutrient of concern was defined as correctly identifying which foods had either 20% or more of the daily value of the nutrient (for the numerical label) or were labeled with “high in” (for the high in labels).

To measure selection of the healthiest food for purchase, the survey asked: “Which of these products would you most want to buy?” The choices were the three foods displayed in random order and participants could only select one food. Selection of the healthiest food for purchase was defined as selecting the food that was high in only one nutrient.

To maximize the realism of the process of reading food labels and selecting products, the survey used a time limit to mirror the typical time people spend making shopping decisions. Research shows that selecting grocery store items takes an average of ∼12 seconds.^39^ However, we lengthened the amount of time given that participants viewed unfamiliar brands and had to scroll through a webpage to look at the products. Pre-testing revealed that the first selection task took the longest to complete, so we used a 30 second time limit for the selection question and 15 seconds for the correct identification questions. For all four outcomes, we recoded missing data as “no” or “incorrect,” including people who had missing data because the timer ran out (4%-10% for all outcomes).

After completing these measures, the survey displayed four types of icon labels in random order: a magnifying glass, an exclamation point in a circle, an exclamation point in an octagon, and an exclamation point in a triangle (stimuli depicted in **Supplementary Figure 1**). The survey asked participants which icon label best signaled when foods are high in sodium, saturated fat, and added sugars and which label most discouraged purchasing of foods high in these nutrients.

### Statistical Analysis

The study design, measures, predictions, and analytic plan were registered before data collection on ClinicalTrials.gov (#NCT06293963). Our pre-registration included a fourth experimental arm consisting of separate icon high-in labels, with one label for each nutrient of concern. However, due to a survey programming error, this arm was not administered, so analyses focus only on the three arms that were administered properly. This error did not impact the randomization of participants in the other three arms.

Analyses were conducted in Stata MP (v.18), with two-tailed tests and a critical alpha of .05. All confidence intervals presented used a 95% confidence level. We predicted that correct identification of the healthiest food would be highest for participants assigned to the icon high-in label, followed by the text high-in label, and then the numerical label. We predicted the same pattern for the other three outcomes. We used the numerical label as the referent because companies already voluntarily display numerical labels on many foods in the US.^30^

We powered the study to detect a difference in the co-primary outcomes between the two label conditions that we anticipated would be most similar: the text and the icon high-in labels. Power analyses revealed that a sample of ∼1,000 per condition yielded 80% power to detect effects of OR=1.16 or larger between any two label conditions for the co-primary outcomes. This is a conservative estimate of effect size based on prior studies showing effect sizes of OR=1.10-1.80 between similar labels.^26,28,29,40^

Consistent with our pre-registration, we excluded participants who completed the survey implausibly quickly (defined as <1/3 of the median completion time, or 4.6 minutes) and those who completed less than 90% of the survey. Other than these exclusions, analyses included all participants according to the experimental condition to which they were randomized (see CONSORT diagram, **Figure 1**). Analyses used mixed effects logistic regression models to examine the impact of experimental condition on the outcomes, accounting for repeated measures within participants. In four separate models, we regressed each outcome on an indicator variable for the experimental condition (excluding the numerical label as the referent) and an indicator variable for food category, treating the intercept as random. Analyses estimated average differential effects for the high-in labels compared to the numerical label, representing the differences in predicted probabilities by label type. We used Chi-square tests with the *margins* command to statistically compare the two high-in labels to each other. We had planned to correct p-values from the additional three pairwise comparisons among high-in label types, but after dropping the fourth experimental arm, we only had one additional pairwise comparison per outcome, so this correction was no longer needed. We then descriptively explored the impact of label type on the primary and secondary outcomes when stratifying by product type.

To examine whether the impact of experimental condition on the co-primary outcomes differed by English proficiency, we regressed each outcome on label type, product type, English proficiency (high vs. limited), and the interaction between label type and English proficiency. These analyses used linear probability models (rather than logistic regression) to aid in interpretation,^41^ as recommended for examining moderation for binary outcomes. Since parents may have heightened awareness of a product’s nutritional content, we used the same approach to determine whether the effects differed by parental status.

Finally, to assess which icon designs are most promising for inclusion in labels, we calculated the proportion of participants that selected each icon label as best signaling when foods are high in sodium, saturated fat, and added sugars and the proportion that selected each label as most discouraging of purchasing foods high in these nutrients.

## Results

The mean age of participants was 35 years, and 50% were women (**Table 1**). About half of participants (49%) had limited English proficiency and about two-thirds (64%) were born in the US. About a third (32%) of participants were of Mexican descent, 15% Spanish, 14% South American, 10% Puerto Rican, 9% Cuban, and 9% Central American (see **Supplementary Table 2** for detailed data on cultural heritage). In terms of racial identity, 56% self-identified as White, 16% as Black, African American, or Afro Latino/a, and 9% as indigenous or American Indian.

**Table 1.** Participant demographics (*n*=3,053)

|  | **Numerical**<br>***n* (%)** | **Text-only**<br>***n* (%)** | **Icon**<br>***n* (%)** | **Total**<br>***n* (%)** |
| --- | --- | --- | --- | --- |
| ***n*** | 1,018 (33%) | 1,008 (33%) | 1,027 (34%) | 3,053 (100%) |
| **Age** |  |  |  |  |
| 18-25 years | 213 (21%) | 186 (18%) | 231 (22%) | 630 (21%) |
| 26-34 years | 294 (29%) | 289 (29%) | 288 (28%) | 871 (29%) |
| 35-44 years | 347 (34%) | 374 (37%) | 347 (34%) | 1,068 (35%) |
| 45-55 years | 164 (16%) | 159 (16%) | 161 (16%) | 484 (16%) |
| Mean, in years (SD) | 35 (9) | 35 (9) | 34 (9) | 35 (9) |
| **Gender identity** |  |  |  |  |
| Woman | 517 (51%) | 493 (49%) | 504 (49%) | 1,514 (50%) |
| Man | 479 (47%) | 496 (49%) | 504 (49%) | 1,479 (49%) |
| Non-binary | 13 (1%) | 16 (2%) | 15 (1%) | 44 (1%) |
| Other | 1 (0%) | 0 (0%) | 1 (0%) | 2 (0%) |
| **Sexual orientation** |  |  |  |  |
| Straight or heterosexual | 897 (91%) | 910 (93%) | 878 (89%) | 2,685 (91%) |
| Gay or lesbian | 39 (4%) | 34 (3%) | 41 (4%) | 114 (4%) |
| Bisexual | 46 (5%) | 30 (3%) | 57 (6%) | 133 (4%) |
| Queer | 5 (1%) | 7 (1%) | 11 (1%) | 23 (1%) |
| Another identity | 1 (0%) | 1 (0%) | 3 (0%) | 5 (0%) |
| **English proficiency** |  |  |  |  |
| Limited | 485 (48%) | 499 (50%) | 514 (50%) | 1,498 (49%) |
| High | 533 (52%) | 509 (50%) | 513 (50%) | 1,555 (51%) |
| **Heritage** |  |  |  |  |
| Mexico | 335 (33%) | 310 (31%) | 321 (31%) | 966 (32%) |
| Spain | 151 (15%) | 153 (15%) | 158 (15%) | 462 (15%) |
| South America | 148 (15%) | 160 (16%) | 128 (13%) | 436 (14%) |
| Puerto Rico | 90 (9%) | 92 (9%) | 108 (11%) | 290 (10%) |
| Cuba | 88 (9%) | 97 (10%) | 97 (10%) | 282 (9%) |
| Central America | 77 (8%) | 86 (9%) | 101 (10%) | 264 (9%) |
| More than one | 78 (8%) | 70 (7%) | 70 (7%) | 218 (7%) |
| Another heritage | 49 (5%) | 39 (4%) | 38 (4%) | 126 (4%) |
| **Race** |  |  |  |  |
| White | 556 (56%) | 578 (58%) | 559 (55%) | 1,693 (56%) |
| Black, African American, or Afro Latino/a | 159 (16%) | 154 (16%) | 170 (17%) | 483 (16%) |
| American Indian | 64 (6%) | 63 (6%) | 63 (6%) | 190 (6%) |
| Indigenous | 35 (4%) | 32 (3%) | 33 (3%) | 100 (3%) |
| Another race | 7 (1%) | 7 (1%) | 7 (1%) | 21 (1%) |
| More than one | 31 (3%) | 29 (3%) | 32 (3%) | 92 (3%) |
| None | 143 (14%) | 129 (13%) | 148 (15%) | 420 (14%) |
| **Born in the US** | 660 (65%) | 638 (63%) | 666 (65%) | 1,964 (64%) |
| **Years lived in the US (among foreign-born)** |  |  |  |  |
| 1 year or less | 61 (6%) | 59 (6%) | 58 (6%) | 178 (17%) |
| 2-4 years | 84 (8%) | 84 (8%) | 91 (9%) | 259 (24%) |
| 5-9 years | 78 (8%) | 97 (10%) | 75 (7%) | 250 (23%) |
| 10-19 years | 60 (6%) | 78 (8%) | 74 (7%) | 212 (20%) |
| 20 or more years | 66 (7%) | 43 (4%) | 56 (5%) | 165 (16%) |
| **Body Mass Index (BMI)** |  |  |  |  |
| Underweight | 124 (12%) | 121 (12%) | 109 (11%) | 354 (12%) |
| Healthy weight | 423 (42%) | 411 (41%) | 444 (44%) | 1,278 (42%) |
| Overweight | 247 (25%) | 261 (26%) | 232 (23%) | 740 (25%) |
| Obesity | 207 (21%) | 204 (20%) | 230 (23%) | 641 (21%) |
| **Diagnosed with pre-diabetes or diabetes** | 141 (14%) | 132 (13%) | 133 (13%) | 406 (13%) |
| **Diagnosed with high blood pressure** | 104 (10%) | 97 (10%) | 107 (11%) | 308 (10%) |
| **Mental health** |  |  |  |  |
| Excellent | 299 (30%) | 325 (32%) | 329 (32%) | 953 (32%) |
| Very good | 348 (35%) | 321 (32%) | 329 (32%) | 998 (33%) |
| Good | 227 (23%) | 235 (23%) | 238 (23%) | 700 (23%) |
| Fair or poor | 133 (13%) | 120 (12%) | 121 (12%) | 374 (12%) |
| **Education level** |  |  |  |  |
| High school degree (GED) or less | 271 (27%) | 245 (24%) | 259 (25%) | 775 (25%) |
| Associate's degree or some college | 359 (35%) | 338 (34%) | 367 (36%) | 1,064 (35%) |
| Bachelor's degree or higher | 388 (38%) | 424 (42%) | 400 (39%) | 1,212 (40%) |
| **Parental status** |  |  |  |  |
| No children | 312 (31%) | 298 (30%) | 306 (30%) | 916 (31%) |
| 1 or more children | 687 (69%) | 691 (70%) | 707 (70%) | 2,085 (69%) |
| **Annual household income** |  |  |  |  |
| \$24,999 or less | 231 (23%) | 220 (22%) | 229 (23%) | 680 (23%) |
| \$25,000 to \$49,999 | 148 (15%) | 160 (16%) | 163 (16%) | 471 (16%) |
| \$50,000 to \$99,999 | 276 (28%) | 271 (27%) | 243 (24%) | 790 (26%) |
| \$100,000 to \$149,999 | 174 (17%) | 165 (17%) | 184 (18%) | 523 (17%) |
| \$150,000 or more | 174 (17%) | 177 (18%) | 194 (19%) | 545 (18%) |
| **Received SNAP in the last 12 months** | 412 (41%) | 431 (43%) | 450 (44%) | 1,293 (43%) |
| **Received WIC in the last 12 months** | 313 (31%) | 324 (33%) | 317 (31%) | 954 (32%) |

*Note.* Percentage of missing values ranged from 0 to 3%.

The text high-in label led to significantly higher correct identification of the least healthy food compared to the numerical label (49% correct vs. 44% correct, *p*=.002, ADE=4.89 percentage points; **Table 2, Supplementary Table 3)**. The icon high-in labels did not lead to significantly higher correct identification of the least healthy food compared to the numerical label (47% correct, *p*=.07). The text high-in and icon high-in labels did not lead to higher identification of the healthiest product compared to numerical labels (all *p*>=.71) or to higher correct identification of foods high in nutrients of concern (all *p*>=.13). Likewise, the text high-in label and the icon high-in label did not lead to higher selection of the healthiest food to purchase (all *p*>=.09). There were no differences between text high-in and icon high-in on any outcomes (all *p*>=.19). The pattern of experimental findings stratified by product type was very similar to results from the mixed models for all three product types (**Supplementary Table 4**).

**Table 2.**
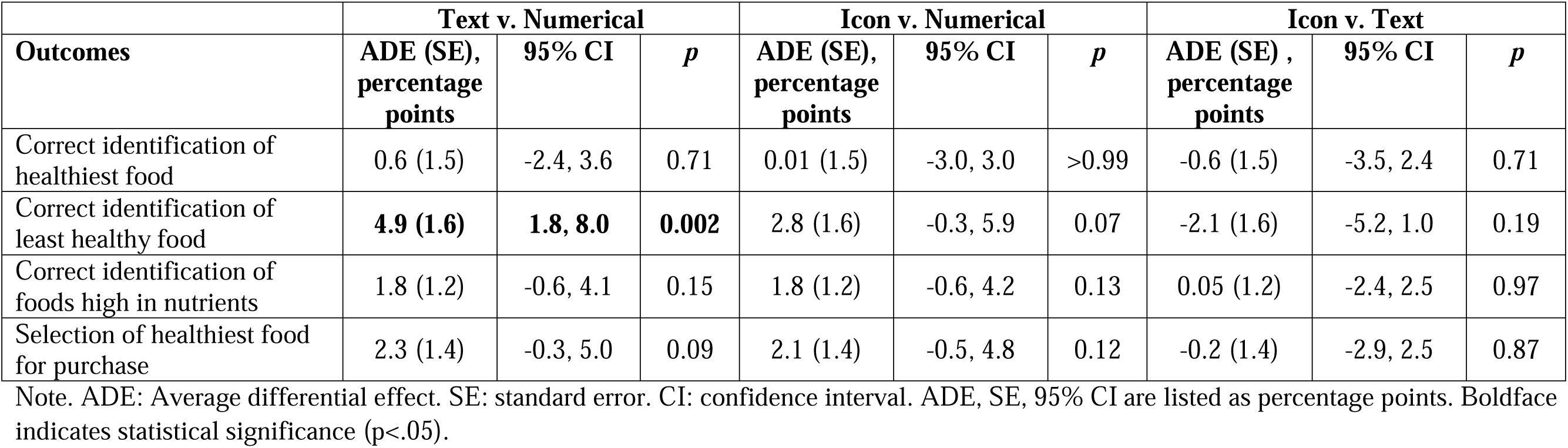
Impact of front-of-package nutrition labels on study outcomes (n = 3,053, n observations = 9,159)

| Outcomes | Text v. Numerical |  |  | Icon v. Numerical |  |  | Icon v. Text |  |  |
| --- | --- | --- | --- | --- | --- | --- | --- | --- | --- |
|  | ADE (SE),<br>percentage<br>points | 95% CI | *p* | ADE (SE),<br>percentage<br>points | 95% CI | *p* | ADE (SE) ,<br>percentage<br>points | 95% CI | *p* |
| Correct identification of healthiest food | 0.6 (1.5) | -2.4, 3.6 | 0.71 | 0.01 (1.5) | -3.0, 3.0 | >0.99 | -0.6 (1.5) | -3.5, 2.4 | 0.71 |
| Correct identification of least healthy food | **4.9 (1.6)** | **1.8, 8.0** | **0.002** | 2.8 (1.6) | -0.3, 5.9 | 0.07 | -2.1 (1.6) | -5.2, 1.0 | 0.19 |
| Correct identification of foods high in nutrients | 1.8 (1.2) | -0.6, 4.1 | 0.15 | 1.8 (1.2) | -0.6, 4.2 | 0.13 | 0.05 (1.2) | -2.4, 2.5 | 0.97 |
| Selection of healthiest food for purchase | 2.3 (1.4) | -0.3, 5.0 | 0.09 | 2.1 (1.4) | -0.5, 4.8 | 0.12 | -0.2 (1.4) | -2.9, 2.5 | 0.87 |

Note. ADE: Average differential effect. SE: standard error. CI: confidence interval. ADE, SE, 95% CI are listed as percentage points. Boldface indicates statistical significance (p<.05).

English proficiency moderated the impact of label type on correct identification of the least healthy food (interaction *p*=.003). The pattern of moderation indicated that there was a benefit of both high-in labels, compared to the numerical labels for participants with high English proficiency (both *p*<.001), with no differences between labels for limited English proficiency (**Figure 2**). Among participants with high English proficiency, 52% in the text high-in label arm and 52% in the icon high-in label arm correctly identified the least healthy food, compared with 44% in the numerical arm. In contrast, among those with limited English proficiency, 46% in the text high-in label arm and 42% in the icon high-in label correctly identified the least healthy food, compared with 44% in the numerical arm. English proficiency did not moderate the impact of label type on ability to identify the healthiest foods (*p* for interaction=.39). Parental status did not moderate the impact of label type on the ability to identify the healthiest (*p* for interaction=.69) or least healthy foods (*p* for interaction=.60).

**Figure 2.**
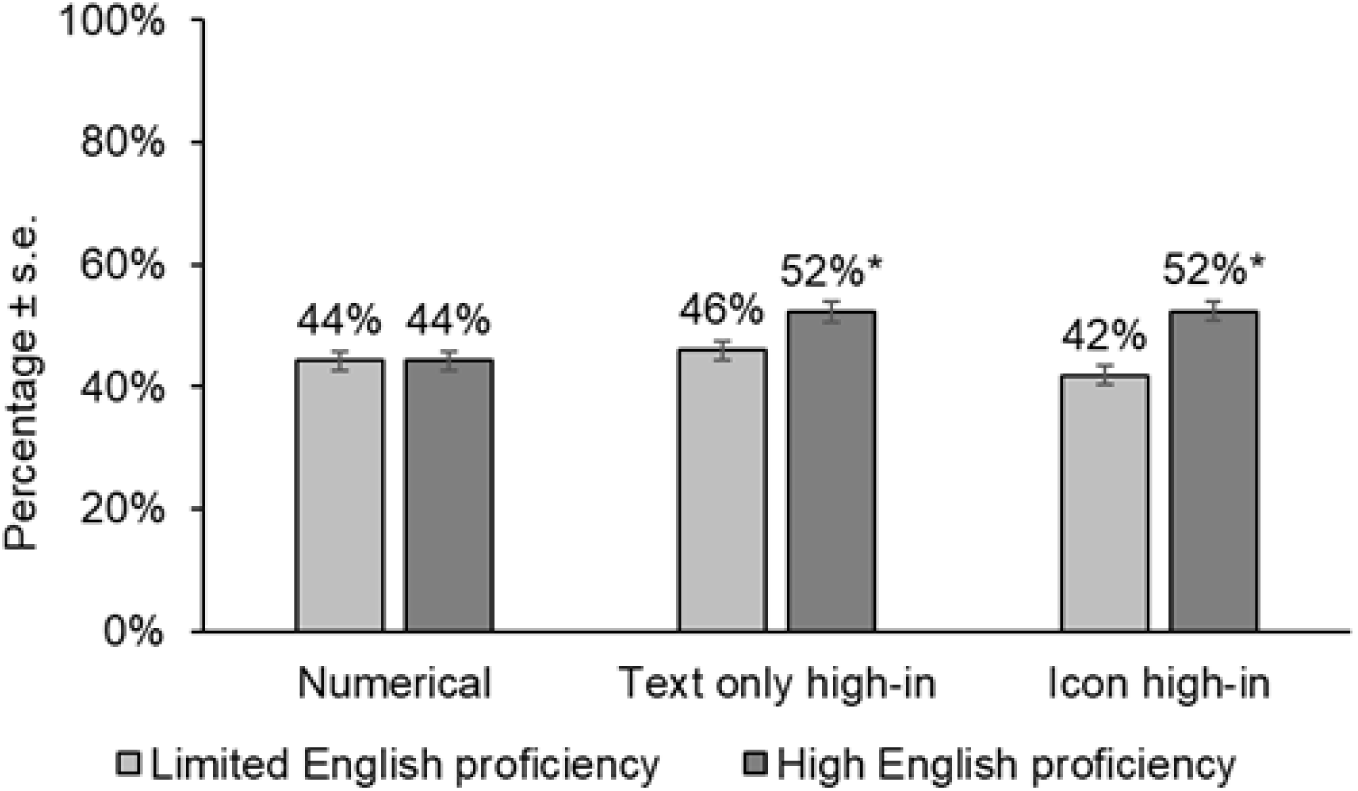
Moderation of English proficiency on the effect of front-of-package nutrition labels for the correct identification of the least healthy foods (*n=*3,053). Limited English proficiency defined as speaking English “not well” or “not at all.” Note: (*) indicates a statistically significant difference (*p*<.001) in the effect between each high-in label v. the numerical label (referent), stratified by English proficiency level, on the correct identification of the least healthy foods.

Finally, when asked which icon best signaled which foods were high in nutrients of concern, 38% of participants selected the triangle, 24% selected the magnifying glass, 20% selected the octagon, and 17% selected the circle (**Supplementary Figure 3, Panel A**). The pattern was similar for which icon most discouraged purchasing of foods high in nutrients of concern, with 41% selecting the triangle, 23% selecting the octagon, 20% selecting the magnifying glass, and 16% selecting the circle (**Supplementary Figure 3, Panel B**).

## Discussion

In this online randomized trial with Latine consumers, we examined the impact of front-of-package nutrition labels signaling when foods were high in nutrients of concern (i.e., high-in labels), compared with numerical labels. The labels were based on designs developed by FDA in its recent rulemaking process. Text high-in labels led to improvements in the ability to identify the least healthy food, compared to numerical labels. There were no differences between label type in the ability to identify the healthiest food, the ability to identify which foods were high in nutrients of concern, or selection of the healthiest food for purchase.

We predicted that high-in labels would perform better than the numerical labels on all outcomes. The current study suggests that text high-in labels may be more effective than numerical labels at helping Latine consumers identify which products are most unhealthy. However, in this study, high-in labels performed comparably to the numerical label on helping consumers identify healthy products, identify products high in nutrients of concern, and select healthier products. These results stand in contrast to prior studies that found that consumers tend to struggle with numerical nutrition information, including the concept of percent daily value^31–34^ as well as quasi-experimental and experimental studies of Latin American-style high-in labels that indicate substantial benefits of interpretative high-in labels.^19,20,24–29^ One possible explanation for why the high-in labels did not outperform numerical labels on all outcomes is that the FDA designs for high-in labels differ substantially from Latin-American-style high-in labels. For example, Latin-American-style labels typically appear in black octagons with white text, while FDA’s designs appear in white rectangles with black text. Additionally, Latin-American-style labels show multiple labels, one label for every nutrient of concern the product is high in, such that consumers can use the number of labels as a heuristic for decision-making,^42^ while FDA’s designs only include one label. Another possible explanation for why the high-in labels did not outperform the numerical labels on some outcomes is that the numerical labels commonly appear voluntarily in the US and in Latin America, so participants were likely already familiar with them. In contrast, FDA’s high-in labels are not used in any countries and would have been novel to participants. Ultimately, the label FDA has proposed to use includes a mix of interpretative information (designating when products are high, medium, or low in nutrients of concerns) and numerical information (percent daily value of nutrients of concern). Although we did not test labels that combine interpretative and numerical information, our results suggest that the way FDA has included interpretative information in its labels may not be as strong as Latin-American-style interpretative high-in labels at informing consumers or encouraging healthier purchases.

Moderation analyses revealed that text high-in label and the same label with an icon led to more accurate identification of unhealthy products compared to numerical labels only for participants with high English proficiency, but not for those with limited English proficiency. It is important to note that we do not know whether English proficiency or other characteristics are driving the moderation effects. These findings were unexpected in light of a prior study that found that that the benefits of icon warnings are even greater in populations with limited (vs high) English proficiency.^35^ One potential explanation for our finding is that the numerical labels were perceived as more credible or believable compared to English-language text among those with limited English proficiency. Prior studies have found that the impact of nutrition labels differs based on English proficiency^35,43^ and language of the warning,^44^ but FDA’s research on its front-of-package labels has not included non-English-speaking participants. In most cases, front-of-package nutrition labels tend to work similarly well across diverse populations,^45–49^ but English proficiency appears to be an important moderator of label effects.^35,43^ Taken together, these findings indicate that that inclusion of meaningful numbers of non-English speaking populations in research on nutrition labels is important for promoting equitable label design and avoiding the potential for labels to exacerbate diet-related disease among the 8% of the US population that has limited English proficiency.^22^

This study also examined which kinds of icons would be best to include in front-of-package nutrition labels. A triangle icon with an exclamation mark was more promising than a magnifying glass, an octagon with an exclamation mark, and a circle with an exclamation mark. In this study, adding a magnifying glass (like the one used in Canada’s and Brazil’s labels) to the high-in labels did not enhance effectiveness. Moreover, participants perceived the triangle icon to be more effective than the magnifying glass icon when pitting them head-to-head, similar to findings in one prior study.^26^ This may be because such an icon is already used on the packaging of US products to caution consumers about a range of hazards; for example, the symbol appears on cancer warnings in California^50^ and alongside DANGER, CAUTION, and WARNING labels recommended by the American National Standards Institute.^51^

### Limitations

Strengths of this study include the participation of many people with limited English proficiency and the use of professionally designed stimuli. One limitation is that we tested labels in an online setting and could not assess actual purchasing or consumption. Future studies should examine whether these findings extend to real-world settings, such as in shopping environments when consumers make quick decisions with other factors at play such as price, marketing, and product availability. Additionally, the use of a convenience sample means the generalizability to other populations has not yet been established. However, experimental findings from convenience samples tend to replicate those from nationally representative samples.^52–54^ We tested a limited set of products with unfamiliar brands and packaging, leaving open the possibility that results would differ if we had used products with different nutritional profiles, products that are frequently consumed in this study population, or products with real brands. In this study, we did not provide any educational information about how to use the labels, which could have limited the benefits of the novel high-in labels. Finally, we only tested products high in at least one nutrient of concern; future studies should examine how labels perform on foods that are not high in any nutrients of concern.

## Conclusions

High-in labels based on FDA’s designs helped Latine consumers identify less healthy foods compared to numerical labels, but these benefits did not extend to those with limited English proficiency. A text high-in label did not lead to higher correct identification of healthier foods, higher correct identification of foods high in nutrients of concern, or greater selection of healthier foods compared to numerical labels. Additionally, adding a magnifying glass icon to the high-in label did not improve effectiveness compared to a text-only label. Given the large body of prior research showing that Latin-American-style high-in label designs tend to communicate information more effectively than numerical labels, our findings suggest that better-designed high-in labels may be needed to support Latine consumers and populations with limited English proficiency in identifying and selecting healthier foods.

## Data Availability

Data may be available upon request to the authors.

## Acknowledgements

We thank Emily Busey and Sara Cathey for designing the study stimuli and Bridget Hollingsworth for reviewing nutrition profiles of study stimuli. We thank Isabella Higgins, Cindy Granados Evans, and Jonathan Lara-Arevalo for providing feedback on Spanish translations of the survey.

## References

1. U.S. Department of Agriculture. Nutrient Intakes from Food and Beverages, by Gender and Age. What We Eat in America, NHANES 2017-March 2020 Prepandemic, https://www.ars.usda.gov/ARSUserFiles/80400530/pdf/1720/Table_1_NIN_GEN_1720.pdf.

2. U.S. Department of Agriculture. Food Pattern Equivalents. What We Eat in America, NHANES 2017-2018 https://www.ars.usda.gov/ARSUserFiles/80400530/pdf/FPED/tables_1-4_FPED_1718.pdf

3. Tsao CW, Aday AW, Almarzooq ZI, et al. Heart Disease and Stroke Statistics—2023 Update: A Report From the American Heart Association. Circulation. 2023;147(8):e93-e621.

4. Fryar CD, Carroll MD, Afful J. Prevalence of overweight, obesity, and severe obesity among adults aged 20 and over: United States, 1960–1962 through 2017–2018. 2020.

5. Guerra ZC, Moore JR, Londoño T, Castro Y. Associations of Acculturation and Gender with Obesity and Physical Activity among Latinos. Am J Health Behav. 2022;46(3):324–336.

6. CDC. National Diabetes Statistics Report, 2020.

7. Aguayo-Mazzucato C, Diaque P, Hernandez S, Rosas S, Kostic A, Caballero AE. Understanding the growing epidemic of type 2 diabetes in the Hispanic population living in the United States. Diabetes Metab Res Rev. 2019;35(2):e3097.

8. Herbert BM, Johnson AE, Paasche-Orlow MK, Brooks MM, Magnani JW. Disparities in Reporting a History of Cardiovascular Disease Among Adults With Limited English Proficiency and Angina. JAMA Netw Open. 2021;4(12):e2138780.

9. Kim EJ, Kim T, Paasche-Orlow MK, Rose AJ, Hanchate AD. Disparities in Hypertension Associated with Limited English Proficiency. J Gen Intern Med. 2017;32(6):632–639.

10. Christoph MJ, Larson N, Laska MN, Neumark-Sztainer D. Nutrition Facts Panels: Who Uses Them, What Do They Use, and How Does Use Relate to Dietary Intake? Journal of the Academy of Nutrition and Dietetics. 2018;118(2):217–228.

11. Hammond D, Acton RB, Rynard VL, et al. Awareness, use and understanding of nutrition labels among children and youth from six countries: findings from the 2019– 2020 International Food Policy Study. International Journal of Behavioral Nutrition and Physical Activity. 2023;20(1):55.

12. Spring 2024 Unified Agenda of Regulatory and Deregulatory Actions. 2024; https://www.reginfo.gov/public/do/eAgendaMain. Accessed August 8, 2024.

13. UNC Global Food Research Program. Countries with mandatory warning labels on packaged foods and drinks. 2024; https://www.globalfoodresearchprogram.org/resource/front-of-package-label-maps/. Accessed September 19, 2024.

14. An R, Liu J, Liu R, Barker AR, Figueroa RB, McBride TD. Impact of Sugar-Sweetened Beverage Warning Labels on Consumer Behaviors: A Systematic Review and Meta-Analysis. Am J Prev Med. 2021;60(1):115–126.

15. Grummon AH, Hall MG. Sugary drink warnings: A meta-analysis of experimental studies. PLoS Med. 2020;17(5):e1003120.

16. Hersey JC, Wohlgenant KC, Arsenault JE, Kosa KM, Muth MK. Effects of front-of-package and shelf nutrition labeling systems on consumers. Nutr Rev. 2013;71(1):1–14.

17. Clarke N, Pechey E, Kosīte D, et al. Impact of Health Warning Labels on Selection and Consumption of Food and Alcohol Products: Systematic Review with Meta-analysis. Health Psychol Rev. 2020:1–39.

18. Song J, Brown MK, Tan M, et al. Impact of color-coded and warning nutrition labelling schemes: A systematic review and network meta-analysis. PLoS Med. 2021;18(10):e1003765.

19. Taillie LS, Bercholz M, Popkin B, Rebolledo N, Reyes M, Corvalán C. Decreases in purchases of energy, sodium, sugar, and saturated fat 3 years after implementation of the Chilean food labeling and marketing law: An interrupted time series analysis. PLoS Med. 2024;21(9):e1004463.

20. Taillie LS, Reyes M, Colchero MA, Popkin B, Corvalan C. An evaluation of Chile’s Law of Food Labeling and Advertising on sugar-sweetened beverage purchases from 2015 to 2017: A before-and-after study. PLoS Med. 2020;17(2):e1003015.

21. US Census Bureau. 2020 Census Statistics Highlight Local Population Changes and Nation’s Racial and Ethnic Diversity. 2021.

22. Dietrich S, Hernandez E. Language use in the United States: 2019. American Community Survey Reports. 2022.

23. US Food and Drug Administration. Daily Value on the Nutrition and Supplement Facts Labels. 2024; https://www.fda.gov/food/nutrition-facts-label/daily-value-nutrition-and-supplement-facts-labels. Accessed September 19, 2024.

24. Fretes G, Corvalán C, Reyes M, et al. Changes in children’s and adolescents’ dietary intake after the implementation of Chile’s law of food labeling, advertising and sales in schools: a longitudinal study. Int J Behav Nutr Phys Act. 2023;20(1):40.

25. Acton RB, Jones AC, Kirkpatrick SI, Roberto CA, Hammond D. Taxes and front-of-package labels improve the healthiness of beverage and snack purchases: a randomized experimental marketplace. Int J Behav Nutr Phys Act. 2019;16(1):46.

26. Goodman S, Vanderlee L, Acton R, Mahamad S, Hammond D. The Impact of Front-of-Package Label Design on Consumer Understanding of Nutrient Amounts. Nutrients. 2018;10(11).

27. Jáuregui A, White CM, Vanderlee L, et al. Impact of front-of-pack labels on the perceived healthfulness of a sweetened fruit drink: a randomised experiment in five countries. Public Health Nutr. 2022;25(4):1094–1104.

28. White-Barrow V, Gomes FS, Eyre S, et al. Effects of front-of-package nutrition labelling systems on understanding and purchase intention in Jamaica: results from a multiarm randomised controlled trial. BMJ open. 2023;13(4):e065620.

29. Mora-Plazas M, Higgins ICA, Gomez LF, et al. Impact of nutrient warning labels on Colombian consumers’ selection and identification of food and drinks high in sugar, sodium, and saturated fat: A randomized controlled trial. PLoS One. 2024;19(6):e0303514.

30. Lim JH, Rishika R, Janakiraman R, Kannan P. Competitive effects of front-of-package nutrition labeling adoption on nutritional quality: Evidence from facts up front–style labels. Journal of Marketing. 2020;84(6):3–21.

31. Bhawra J, Kirkpatrick SI, Hall MG, Vanderlee L, Thrasher JF, Hammond D. Correlates of Self-Reported and Functional Understanding of Nutrition Labels across 5 Countries in the 2018 International Food Policy Study. J Nutr. 2022;152(Suppl 1):13s–24s.

32. Persoskie A, Hennessy E, Nelson WL. US Consumers’ Understanding of Nutrition Labels in 2013: The Importance of Health Literacy. Prev Chronic Dis. 2017;14:E86.

33. Rothman RL, Housam R, Weiss H, et al. Patient understanding of food labels: the role of literacy and numeracy. Am J Prev Med. 2006;31(5):391–398.

34. U.S. Food and Drug Administration. FSANS Explorer: FDA’s Food Safety and Nutrition Survey (FSANS). . 2019; https://fsans-explorer.fda.gov/. Accessed November 22, 2024.

35. Hall MG, Lazard AJ, Grummon AH, et al. Designing warnings for sugary drinks: A randomized experiment with Latino parents and non-Latino parents. Prev Med. 2021;148:106562.

36. US Food and Drug Administration. Agency Information Collection Activities; Proposed Collection; Comment Request; Quantitative Research on Front of Package Labeling on Packaged Foods. 2023; https://www.federalregister.gov/documents/2023/01/26/2023-01551/agency-information-collection-activities-proposed-collection-comment-request-quantitative-research. Accessed September 19, 2024.

37. Global Food Research Program. Sugary drink taxes around the world. 2023; https://www.globalfoodresearchprogram.org/wp-content/uploads/2023/06/GFRP-UNC_Tax_maps_beverages_2023_06.pdf. Accessed June 2023, 2023.

38. Lazard AJ, Mackert MS, Bock MA, Love B, Dudo A, Atkinson L. Visual assertions: Effects of photo manipulation and dual processing for food advertisements. Visual Communication Quarterly. 2018;25(1):16–30.

39. Anesbury Z, Nenycz Thiel M, Dawes J, Kennedy R. How do shoppers behave online? An observational study of online grocery shopping. Journal of Consumer Behaviour. 2016;15(3):261–270.

40. Khandpur N, Amaral Mais L, Bortoletto Martins AP. A comparative assessment of two different front-of-package nutrition label designs: a randomized experiment in Brazil. PLoS One. 2022;17(4):e0265990.

41. Karaca-Mandic P, Norton EC, Dowd B. Interaction terms in nonlinear models. Health Serv Res. 2012;47(1 Pt 1):255–274.

42. Batis C, Aburto TC, Pedraza LS, et al. Self-reported reactions to the front-of-package warning labelling in Mexico among parents of school-aged children. [pre-print]. 2023.

43. Hall MG, Grummon AH, Whitesell C, et al. Evaluating text, icon, and graphic nutrition labels: An eye tracking experiment with Latino adults in the US. Appetite. 2024;204:107745.

44. Razzouk J, Bilić A, Wackowski OA, Cornacchione Ross J, King Jensen JL. Does warning language impact perceptions? Results from an exploratory experiment comparing English, Spanish, and Dual language E-Cigarette warnings among Spanish speakers in the US. Prev Med Rep. 2021;24:101656.

45. Grummon AH, Reimold AE, Hall MG. Influence of the San Francisco, CA, sugar-sweetened beverage health warning on consumer reactions: Implications for equity from a randomized experiment. J Acad Nutr Diet. 2022;122(2):363–370.e366.

46. Roberto CA, Wong D, Musicus A, Hammond D. The influence of sugar-sweetened beverage health warning labels on parents’ choices. Pediatrics. 2016;137(2):e20153185.

47. Grummon AH, Taillie LS, Golden SD, Hall MG, Ranney LM, Brewer NT. Sugar-sweetened beverage health warnings and purchases: A randomized controlled trial. Am J Prev Med. 2019;57(5):601–610.

48. Acton RB, Kirkpatrick SI, Hammond D. Exploring the main and moderating effects of individual-level characteristics on consumer responses to sugar taxes and front-of-pack nutrition labels in an experimental marketplace. Can J Public Health. 2021.

49. VanEpps EM, Roberto CA. The influence of sugar-sweetened beverage warnings: A randomized trial of adolescents’ choices and beliefs. Am J Prev Med. 2016;51(5):664–672.

50. Proposition 65 Warning Symbol. 2025; https://www.p65warnings.ca.gov/warning-symbol. Accessed February 3, 2025.

51. American National Standards Institute. ANSI Z535.3-2022: Criteria for Safety Symbols. 2022; https://blog.ansi.org/ansi-z535-3-2022-criteria-safety-symbols/. Accessed February 3, 2025.

52. Jeong M, Zhang D, Morgan JC, et al. Similarities and Differences in Tobacco Control Research Findings From Convenience and Probability Samples. Ann Behav Med. 2019;53(5):476–485.

53. Berinsky AJ, Huber GA, Lenz GS. Evaluating online labor markets for experimental research: Amazon.com’s Mechanical Turk. Polit Anal. 2012;20(3):351–368.

54. Weinberg JD, Freese J, McElhattan D. Comparing data characteristics and results of an online factorial survey between a population-based and a crowdsource-recruited sample. Sociol Sci. 2014;1:292–310.

