## Supplementary Materials for "Effects of front-of-package nutrition labels in Latine and limited English proficiency populations: A randomized trial"

**Supplementary Table 1. Survey measures**

| Construct | Item |
|---|---|
| *Outcomes* | |
| Identification of healthiest food | Which of these products is the <u>healthiest</u>? |
| Identification of least healthy food | Which of these products is the <u>least healthy</u>? |
| Identification of foods high in nutrients of concern | Which of these products are high in [nutrient]? Check all that apply. |
| Selection for purchase | Which of these products would you most want to buy? |
| Icon PME* - Signaling | Which of these labels <u>best signals</u> when foods have high amounts of sodium, saturated fat, or added sugars? |
| Icon PME* - Discouragement | Which of these labels would <u>most discourage</u> you from wanting to buy foods that have high amounts of sodium, saturated fat, or added sugars? |
| *Demographics* | |
| Age | How old are you (in years)?<br>Numeric free response |
| Gender identity | Select the option that best describes your gender.<br>Response options: Woman; Man; Nonbinary; Prefer to self-describe |
| Sexual orientation | Which of the following best describes you?<br>Response options: Straight or heterosexual; Gay; Lesbian; Bisexual; Queer; Prefer to self-describe |
| English proficiency | How well do you speak English?<br>Response options: Very well (1) - Not at all (4) |
| Heritage | Which of the following describes your cultural heritage? Check all that apply.<br>Response options: Brazilian; Colombian; Cuban; Dominican; Ecuadorian; Guatemalan; Honduran; Mexican; Peruvian; Puerto Rican; Salvadoran; Spanish/Spaniard; Venezuelan; Another origin (please describe) |
| Race | In addition to being of Hispanic/Latino heritage, which of the following categories would you use to describe yourself? Check all that apply.<br>Response options: Black, African American, or Afro Latino/a; Indigenous; American Indian; White; Another race (please describe); None of these |
| Born in the US | Were you born in the United States?<br>Response options: Yes; No |
| Years lived in the US | For how many years have you lived in the United States?<br>Numeric free response |
| Diagnoses | Have you ever been diagnosed with any of the following by a health professional? Check all that apply.<br>Response options: Pre-diabetes; Type 2 diabetes; Obesity; High blood pressure or hypertension; None of the above |
| Mental health | In general, would you say that your mental health is…<br>Response options: Excellent (1) - Poor (5) |
| Education level | What is the highest level of school you have completed?<br>Response options: Less than high school; High school graduate (or GED); Some college or technical school; Associate’s degree; Bachelor’s degree; Graduate or professional degree |
| Annual household income | Which of the following categories best describes your total household income in the last 12 months?<br>Response options: Less than $10,000; $10,000 to $14,999; $15,000 to $24,999; $25,000 to $34,999; $35,000 to $49,999; $50,000 to $74,999; $75,000 to $99,999; $100,000 to $149,999; $150,000 to $199,999; $200,000 or more |

| | |
|---|---|
| Received SNAP in the last 12 months | In the last 12 months, did you or anyone in your household get SNAP or Food Stamps, even if only for one month?<br>Response options: Yes; No |
| Received WIC in the last 12 months | In the last 12 months, did you or anyone in your house receive WIC, even if only for one month?<br>Response options: Yes; No |

*PME=perceived message effectiveness

**Supplementary Table 2. Cultural heritage by country of origin (n=3,053)**

| **Cultural heritage** | ***n*** (%) |
|---|---|
| Mexican | 966 (32%) |
| Spanish/Spaniard | 462 (15%) |
| Puerto Rican | 290 (10%) |
| Cuban | 282 (9%) |
| Venezuelan | 142 (5%) |
| Colombian | 127 (4%) |
| Dominican | 114 (4%) |
| Salvadoran | 85 (3%) |
| Honduran | 80 (3%) |
| Ecuadorian | 69 (2%) |
| Guatemalan | 65 (2%) |
| Brazilian | 37 (1%) |
| Peruvian | 37 (1%) |
| Nicaraguan | 24 (1%) |
| Argentinian | 8 (0.3%) |
| Chilean | 7 (0.2%) |
| Panamanian | 7 (0.2%) |
| Uruguayan | 5 (0.2%) |
| Costa Rican | 3 (0.1%) |
| Bolivian | 2 (0.1%) |
| Paraguayan | 2 (0.1%) |
| More than one | 218 (7%) |
| Other | 12 (0.4%) |

*Note*: Percentage of missing values was 0.3%.

**Supplementary Table 3. Proportion of participants correctly identifying the healthiest food, least healthy food, foods high in nutrients, and selecting the healthiest food for purchase, by label type (n = 3,053)**

| | Numerical | | Text-only high-in label | | Icon high-in label | |
|---|---|---|---|---|---|---|
| **Outcomes** | **N (%)** | **95% CI** | **N (%)** | **95% CI** | **N (%)** | **95% CI** |
| Correct identification of healthiest food | 1,388 (45%) | 44%, 47% | 1,393 (46%) | 44%, 48% | 1,402 (46%) | 44%, 47% |
| Correct identification of least healthy food | 1,352 (44%) | 43%, 46% | 1,485 (49%) | 47%, 51% | 1,451 (47%) | 45%, 49% |
| Correct identification of foods high in nutrients | 574 (19%) | 17%, 20% | 629 (21%) | 19%, 22% | 640 (21%) | 19%, 22% |
| Selection of healthiest food for purchase | 1,131 (37%) | 35%, 39% | 1,191 (39%) | 38%, 41% | 1,207 (39%) | 37%, 41% |

CI = Confidence Interval

**Supplementary Table 4. Impact of front-of-package nutrition labels on study outcomes stratified by product type (n = 3,053, n observations = 9,159)**

| Outcomes | Product type | Text-only high-in vs Numerical | | | Icon high-in vs Numerical | | | Icon high-in vs Text-only high-in | | |
|---|---|---|---|---|---|---|---|---|---|---|
| | | ADE (SE) | 95% CI | *p* | ADE (SE) | 95% CI | *p* | ADE (SE) | 95% CI | *p* |
| Correct identification of healthiest food | Pizza | 1.12 (2.20) | -3.20, 5.44 | .61 | 2.74 (2.20) | -1.56, 7.04 | .21 | 1.62 (2.20) | -2.70, 5.94 | .46 |
| | Pie | 2.51 (2.21) | -1.81, 6.84 | .26 | 2.84 (2.20) | -1.47, 7.14 | .20 | .32 (2.21) | -4.01, 4.65 | .88 |
| | Chicken | -1.78 (2.22) | -6.14, 2.57 | .42 | **-5.41 (2.21)** | **-9.73, -1.08** | **.01** | -3.63 (2.21) | -7.96, .71 | .10 |
| Correct identification of least healthy food | Pizza | 3.81 (2.21) | -.53, 8.14 | .09 | 3.32 (2.20) | -1.00, 7.63 | .13 | -.49 (2.21) | -4.83, 3.85 | .82 |
| | Pie | **6.86 (2.20)** | **2.54, 11.18** | **.002** | 2.66 (2.19) | -1.63, 6.94 | .22 | -4.20 (2.21) | -8.53, .13 | .06 |
| | Chicken | 3.85 (2.22) | -.51, 8.20 | .08 | 2.50 (2.21) | -1.83, 6.84 | .26 | -1.34 (2.22) | -5.69, 3.00 | .54 |
| Correct identification of foods high in nutrients | Pizza | 1.97 (1.75) | -1.46, 5.39 | .26 | 3.15 (1.76) | -.30, 6.60 | .07 | 1.19 (1.80) | -2.34, 4.71 | .51 |
| | Pie | 2.76 (1.77) | -.70, 6.23 | .12 | .33 (1.72) | -3.04, 3.69 | .85 | -2.44 (1.77) | -5.91, 1.03 | .17 |
| | Chicken | 1.29 (1.79) | -2.23, 4.80 | .47 | 2.46 (1.80) | -1.08, 5.99 | .17 | 1.17 (1.83) | -2.42, 4.75 | .52 |
| Selection of healthiest food for purchase | Pizza | 1.74 (2.13) | -2.44, 5.91 | .42 | .18 (2.11) | -3.96, 4.32 | .93 | -1.56 (2.13) | -5.72, 2.61 | .46 |
| | Pie | **4.47 (2.20)** | **.17, 8.77** | **.04** | 4.22 (2.18) | -.06, 8.50 | .05 | -.25 (2.20) | -4.57, 4.08 | .91 |
| | Chicken | .85 (2.14) | -3.33, 5.04 | .69 | 2.02 (2.13) | -2.16, 6.20 | .34 | 1.17 (2.14) | -3.03, 5.37 | .58 |

Note. ADE: Average differential effect. SE: standard error. CI: confidence interval. ADE, SE, and 95% CI are listed as percentage points. Boldface indicates statistical significance ($p<.05$).

**Supplementary Figure 1. Stimuli used in experiment**

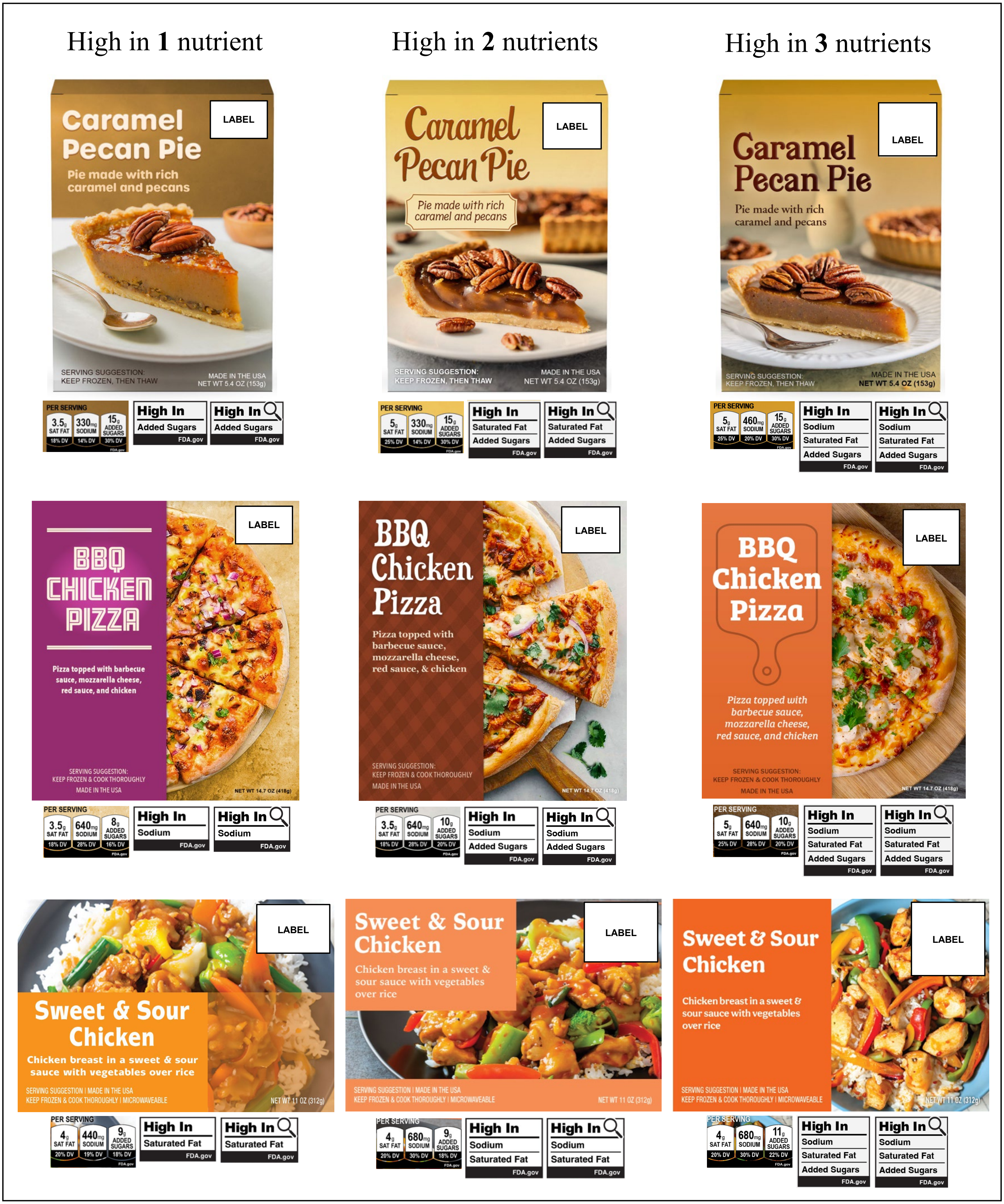

**Supplementary Figure 2. Images used in survey questions about icons**

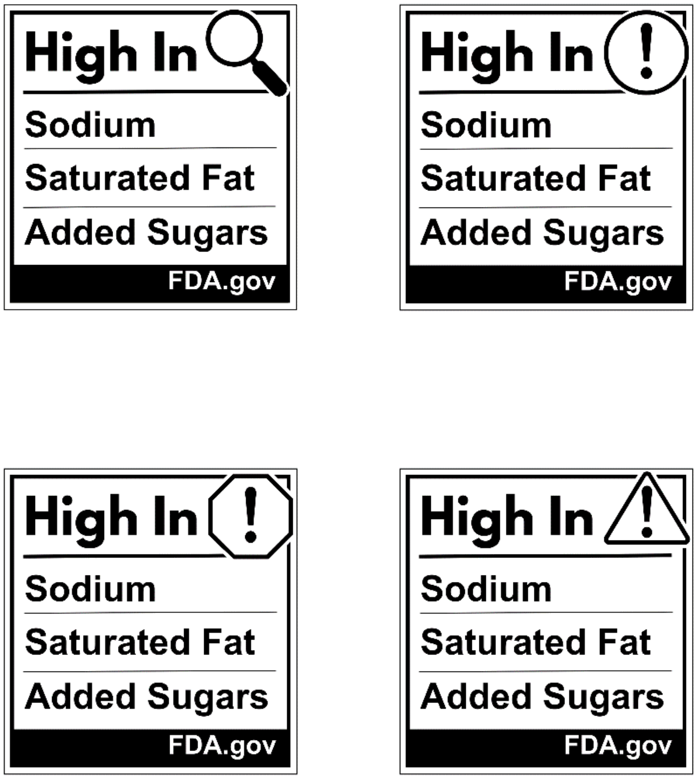

**Supplementary Figure 3. Selection of icons among Latine consumers in the US (Panel A: n=3,048, Panel B: n=3,052). Error bars are standard errors.**

**Panel A**. Proportion of participants selecting each icon as the one that best signals foods with high amounts of nutrients of concern

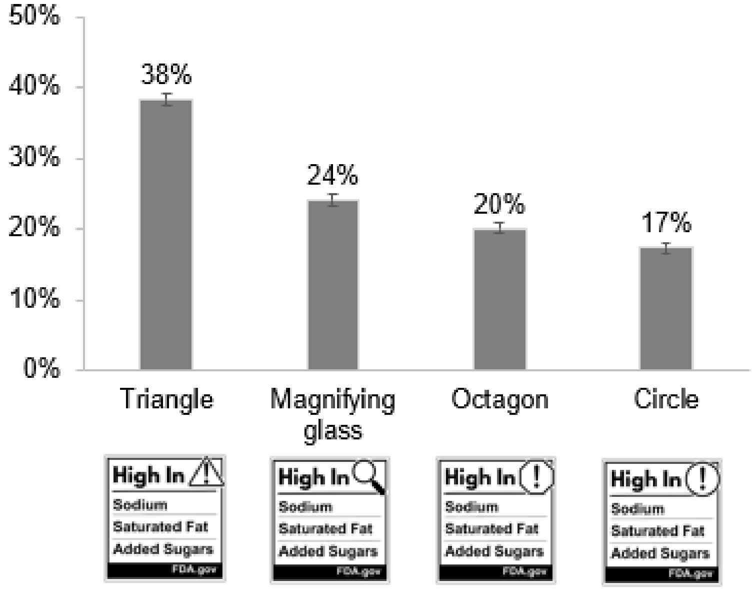

**Panel B**. Proportion of participants selecting each icon as the one that most discourages purchase of foods with high amounts of nutrients of concern

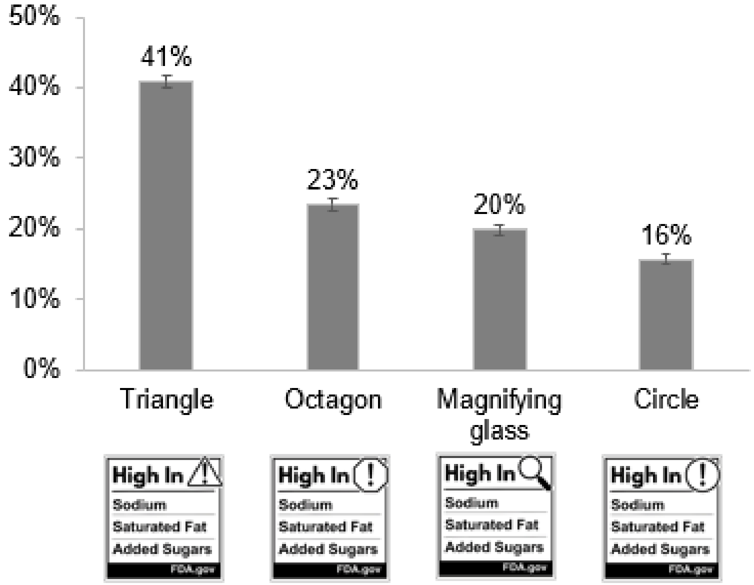
